# Detection of SARS-CoV-2 infection in gargle, spit and sputum specimens

**DOI:** 10.1101/2021.05.02.21255857

**Authors:** Eero Poukka, Henna Mäkelä, Lotta Hagberg, Thuan Vo, Hanna Nohynek, Niina Ikonen, Kirsi Liitsola, Otto Helve, Carita Savolainen-Kopra, Timothée Dub

## Abstract

The gold standard for SARS-CoV-2 infection diagnosis is RT-PCR from nasopharyngeal specimen (NPS). Its collection involves a close contact between patients and healthcare workers requiring a significant amount of workforce and putting them at risk of infection. We evaluated self-collection of alternative specimens and compared their sensitivity and Ct values to NPS. We visited acute COVID-19 outpatients to collect concomitant nasopharyngeal and gargle specimens and had patients self-collect a gargle and either sputum or spit specimens on the next morning.

We included 40 patients and collected 40 concomitant nasopharyngeal and gargle specimens, as well as 40 gargle, 22 spit and 16 sputum specimens on the next day, as 2 patients could not produce sputum.

All specimens were as sensitive as NPS. Gargle specimens had a sensitivity of 0.97 (CI 95% 0.92-1,00), whether collected concomitantly to NPS or on the next morning. Next morning spit and sputum specimens showed a sensitivity of 1.00 CI (95% 1.00-1.00) and 0.94 (CI 95% 0.87-1.00), respectively. The gargle specimens had a significantly higher mean cycle threshold (Ct) values, 29.89 (SD 4.63) (p-value <0.001) and 29.25 (SD 3.99) (p-value <0.001) when collected concomitantly and on the next morning compared to NPS (22.07, SD 4.63). Ct value obtained with spit (23.51, SD 4.57, p-value 0.11) and sputum (25.82, SD 9.21, p-value 0.28) specimens were close to NPS.

All alternative specimen collection methods were as sensitive as NPS, but spit collection appeared more promising, with a low Ct value and ease of collection. Our findings warrant further investigation.

## Introduction

In December 2019, SARS-CoV-2 emerged from the Chinese city of Wuhan (1). The disease spread into countries outside China and was declared a global pandemic in March 2020 (2). Within a year, more than 115 million COVID-19 cases were confirmed, including two and a half million deaths (3).

The SARS-CoV-2 pandemic control relies on a test-trace-isolate strategy with early diagnosis and isolation of infected individuals and identification of their contacts (4) which has led to an initial shortage of personal protective and sampling equipment, as well as increasing healthcare workers’ workload (6,7). Alone in Finland, a country with approximately 5 500 000 inhabitants, by the 20^th^ of March 2021 over 3 700 000 tests, of which 145 000 during the first week of March 2021, were conducted and analyzed nationwide during the COVID-19 pandemic (5,6).

Collection of nasopharyngeal specimen (NPS) is the gold standard for SARS-CoV-2 infection diagnosis (7). However, it is an unpleasant procedure requiring a close contact with HCW with a risk of discomfort and in worst cases, epistaxis for the patient, and infections exposure for the HCW (8,9). The use of an alternative specimen collection method could increase specimen collection and testing capacities as well as decrease HCWs’ workload and risk for infection (8). There has been several studies evaluating alternative specimen collection methods but none of the specimen collection methods have yet superseded NPS (10–13), even though one Finnish private healthcare provider now offers asymptomatic patients the possibility to self-collect a gargle specimen as an alternative to NPS (14).

We evaluated and compared three alternative specimen collection methods that would not require close contact to a HCW and compared their sensitivity and Ct values to NPS.

## Materials and methods

We contacted confirmed COVID-19 outpatients who had been diagnosed with SARS-CoV-2 infection by RT-PCR (reverse transcription polymerase chain reaction) on NPS a few days earlier. Children under 2 years old were not eligible for participation. After calling the patient for recruitment, we visited them on the same and following day.

During the first home visit, we gathered informed consent, gave participants a link to an online symptom-questionnaire and collected a NPS and gargle specimen (gargle 1). We also gave them instructions and containers for collection of gargle (gargle 2) and, depending on the recruitment week, sputum or spit specimen on the next morning. On the following day, the patients collected themselves the second gargle (gargle 2) and either spit or sputum specimens. The next day the self-collected specimens were collected. The timeline of the study is presented in Figure 1.

**Figure 1.**
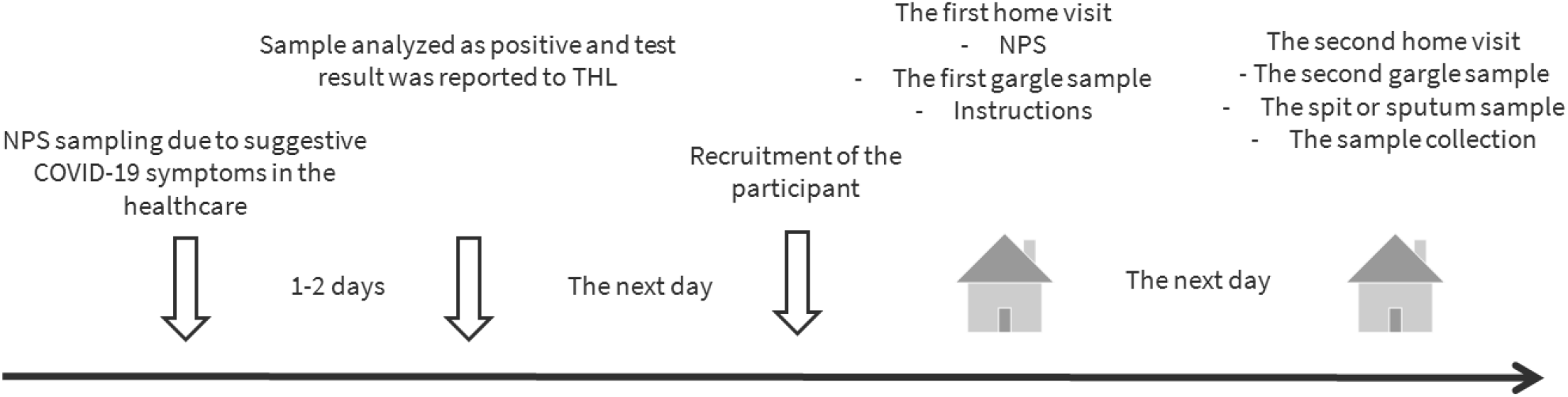
Timeline of the study. NPS =Nasopharyngeal specimen.

**Figure 2.**
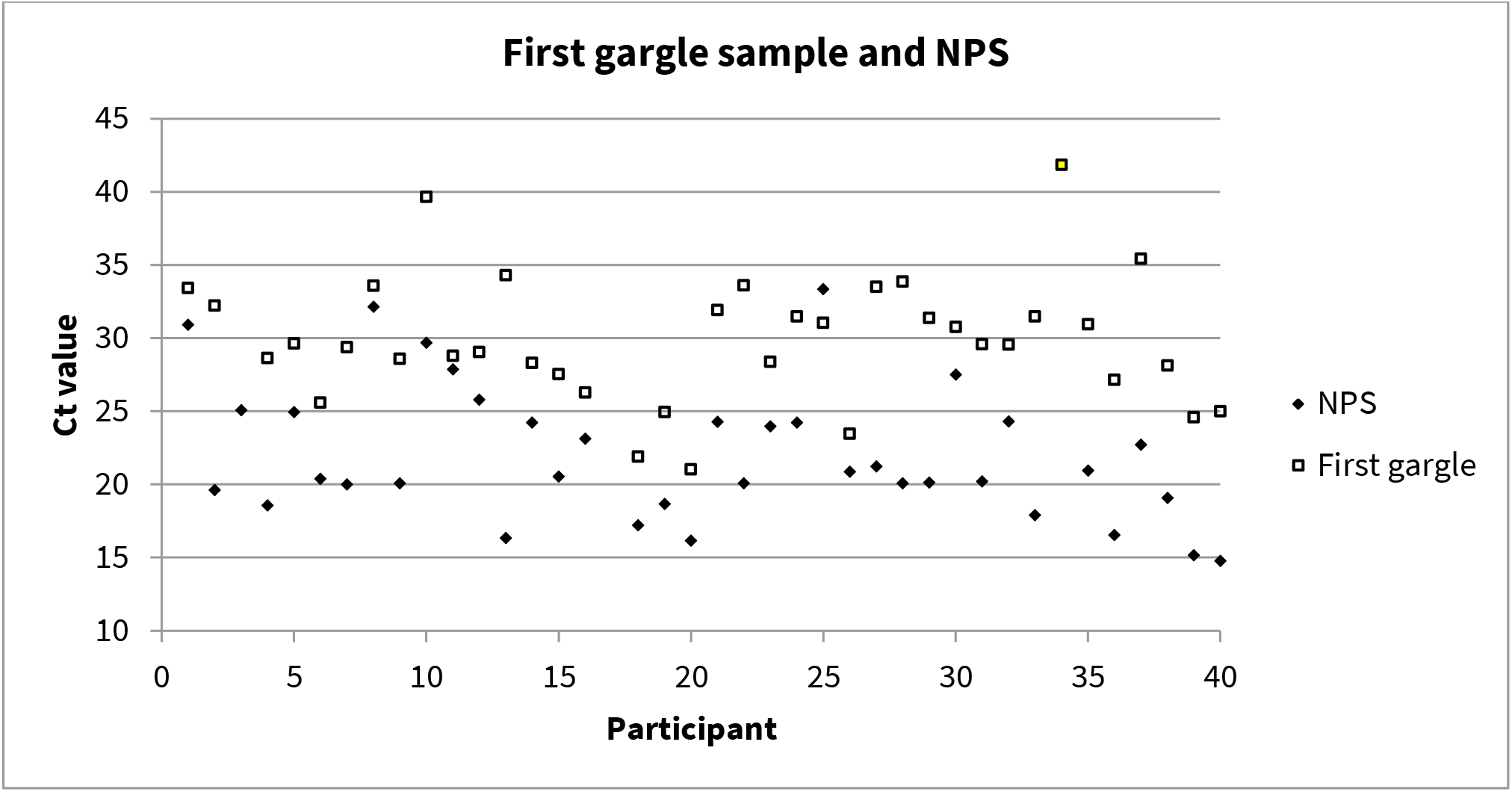
Ct-values from gargle 1 and nasopharyngeal sample by each participant.

**Figure 3.**
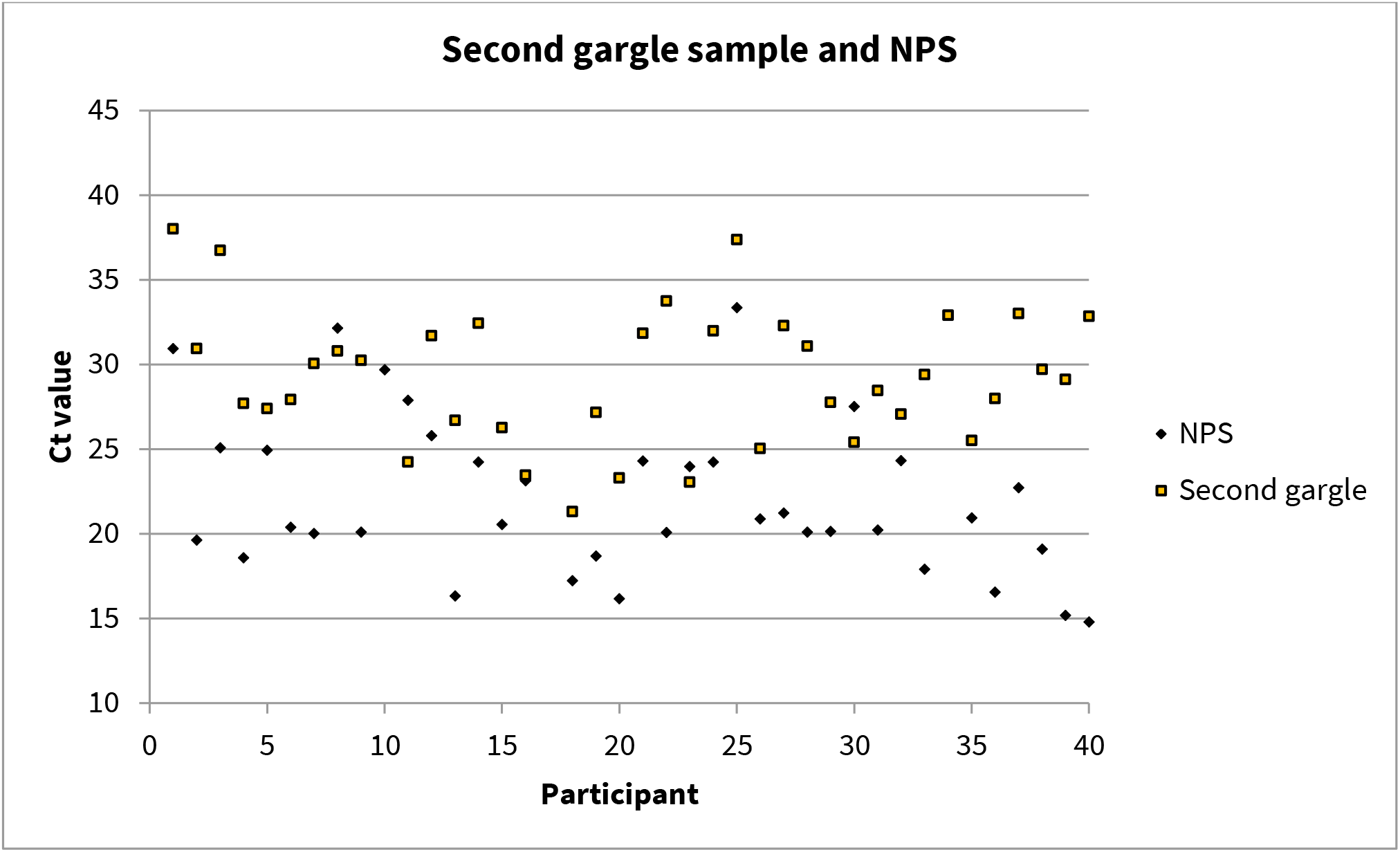
Ct-values from gargle 2 and nasopharyngeal sample by each participant.

**Figure 4.**
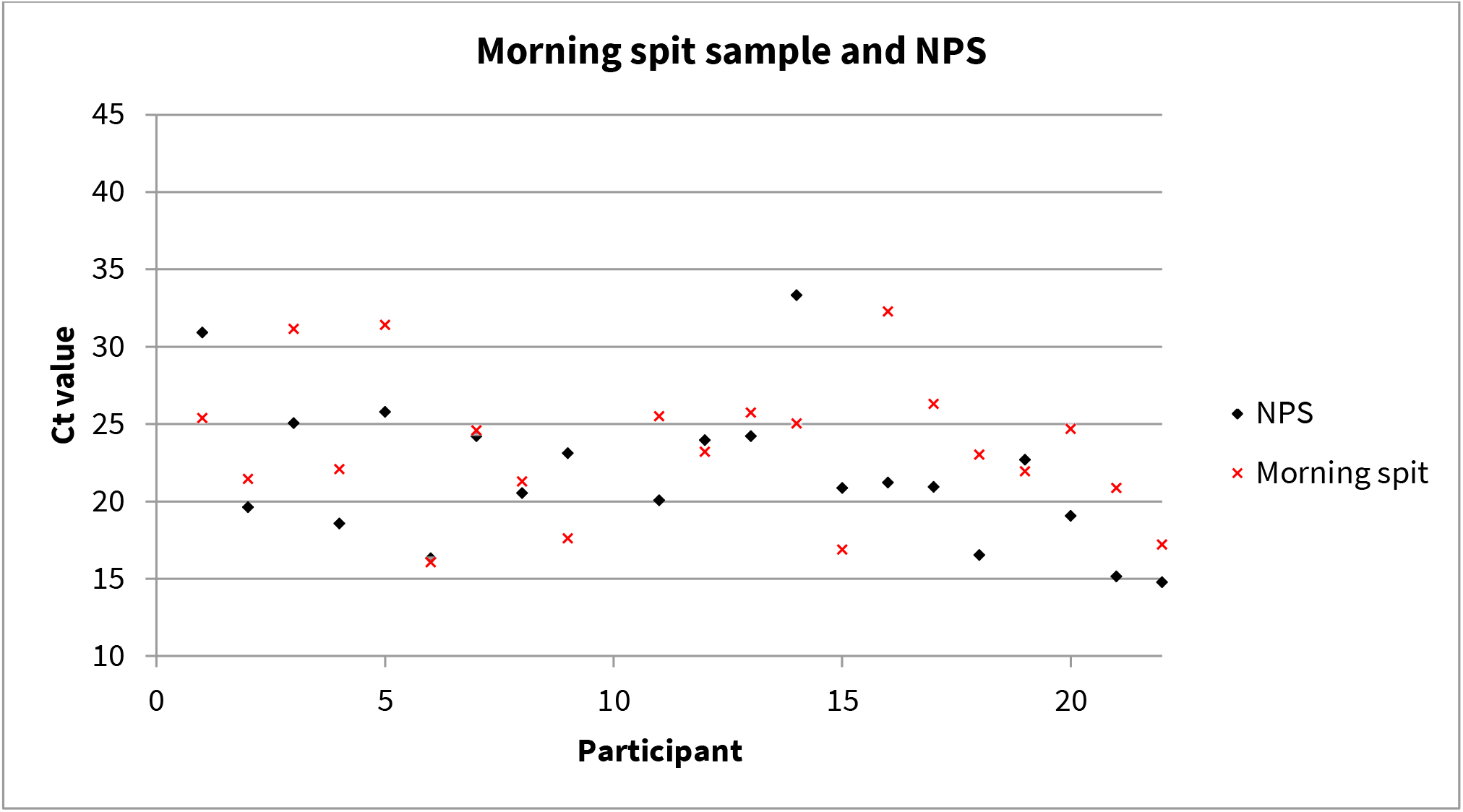
Ct- values from spit sample and nasopharyngeal sample by each participant.

**Figure 5.**
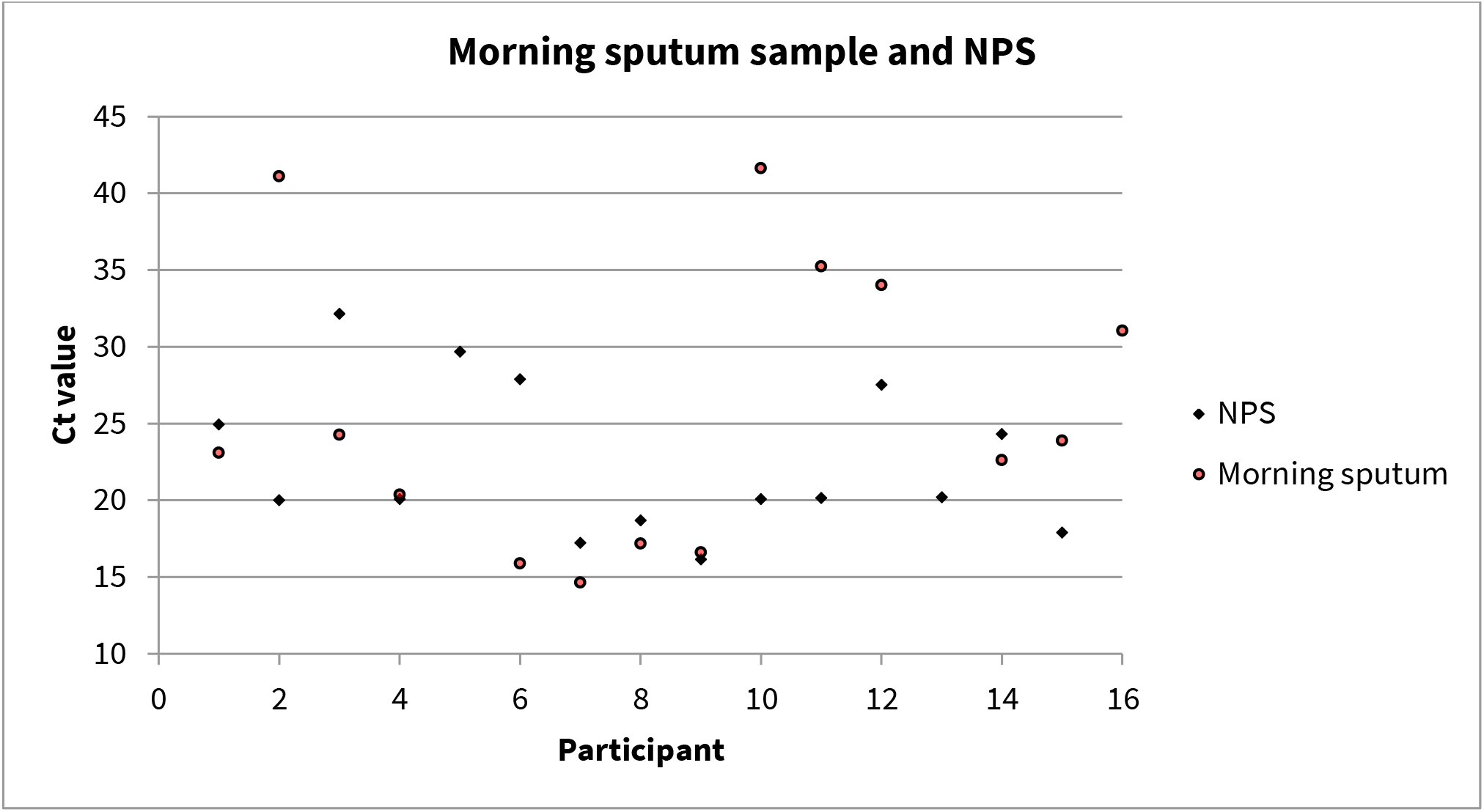
Ct-values from sputum sample and nasopharyngeal sample by each participant.

All alternative specimens were collected into a 70 ml plastic container. To collect the gargle specimens, the patients were asked to have sip of water, and gargle it for 5-20 seconds before spitting it in the container. For spit collection, the patients were asked to spit continuously into the container until filled to half its volume, and with sputum specimen they were asked to cough sputum deep from their lungs and then spit it into the container. Both the spit and sputum specimens were advised to be collected in the morning before the patients ate, drunk or brushed their teeth.

All specimens were transported at room temperature and analyzed on the same day in Expert Microbiology unit at the Finnish Institute for Health and Welfare (THL).

The Finnish communicable diseases law and the law on the duties of THL allows the implementation of this noninvasive research without seeking further ethical approval (THL-laki 5 e §; Tartuntatautilaki 7 § and 23 §).

### Laboratory methods

RNA extraction from samples was performed using Chemagic Viral300 DNA/RNA Kit H96 (PerkinElmer) according to the manufacturer’s instructions. A sample volume 300 μl and an elution volume 50 μl were used. Highly viscose gargle samples were vortexed with 1 ml PBS before taking 300 μl for RNA extraction. Real time RT PCR was performed using qScript™ XLT One-Step RT-qPCR ToughMix® (Quantabio). SARS-CoV-2 was detected using the E (envelope) gene real-time RT-PCR assay. Primers and probes were based on the Corman E gene premier/probe set(15). The thermal profile for PCR was 55°C for 20 min and 95°C for 3 min, followed by 45 cycles at 95°C 15 s and 58 °C 1 min using CFX thermal cycler (BioRad).

### Statistical analysis

We used NPS RT-PCR test results as reference method and Ct values as surrogates for viral load analysis. We performed paired t-test or Wilcoxon signed-rank test to compare the measured Ct values between nasopharyngeal and gargle, and between nasopharyngeal and sputum/spit specimens. Standard methods were used to calculate sensitivity and specificity of the other diagnostic tests (index tests) from saliva- and sputum/spit specimens. Exact McNemar’s test was used to assess the differences in sensitivity and specificity.

We calculated Cohen’s kappa coefficient to evaluate the agreement between the reference and the index tests. Area under a receiver operating characteristic curve (AUC) and 95% CI were reported. Because of the imperfect reference test, latent class analysis was used as a correction method to adjust the estimated sensitivity and specificity of the index tests based on the existing sensitivity and specificity of the reference test. Model selection was based on Bayesian information criterion (BIC). Data analysis was performed using R software (version. 3.6.0).

## Results

We enrolled 40 patients with a mean age of 38.7 years old (SD: 12.6) including 21 (53%) females. Enrolment was done as soon as they received the positive testing results, either one day (n=27/40, 67.5%) or two days (n=13/40, 32.5%) after they had been NPS sampled for COVID-19 diagnosis. Thirty-one patients had been symptomatic since disease onset (supplementary table 1) with most prevalent symptoms being fatigue (86%), headache (81%) and cough (79%). At the time of specimen collection, only 24 patients were symptomatic with the most prevalent being cough (44%), anosmia (42%) and headache (40%).

We collected 40 concomitant nasopharyngeal and gargle specimens on recruitment day, as well as 40 next morning gargle specimens. Out of 22 patients assigned to the spit specimen collection group, all of them gave back specimens, while out of 18 patients assigned to the sputum group, two patients could not produce sputum.

All specimens were generally as sensitive as NPS to diagnose SARS-CoV-2 infection in our study population. The spit specimen showed the highest sensitivity (sensitivity 1.00 CI 95 % 1.00-1.00), followed by gargle specimens, regardless of when they were collected: 0.97 (CI 95 % 0.92-1,00). The sputum specimen had the lowest sensitivity (0.94 CI 95 % 0.87-1.00).

We compared Ct values obtained from alternative specimens to NPS (Table 2). NPS had the lowest Ct value (22.07, SD 4.63), although it was not significantly lower compared to sputum (25.82, SD 9.21), (p-value 0.28) and spit (23.51, SD 4.57), (p-value 0.11) specimens. Both gargle specimens had statistically significantly higher Ct values, compared to NPS (Table 2).

**Table 1.**
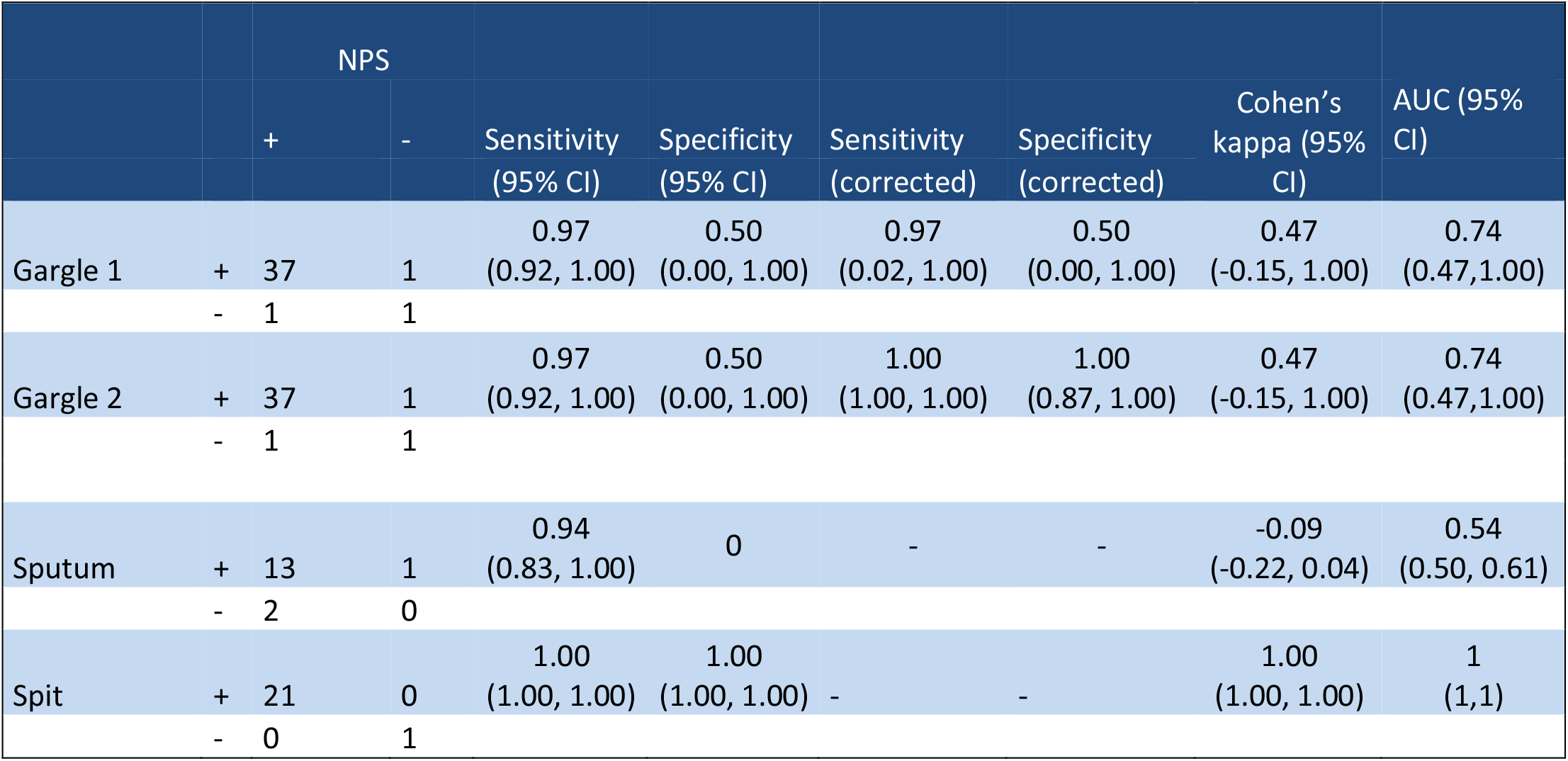
Sensitivity and specificity of tested sampling methods

**Table 2:** Comparison of Ct-values obtained with alternative samples to nasopharyngeal samples.

| Sample | n | Mean (SD)/ | p-value |
| --- | --- | --- | --- |
| Nasopharyngeal | 40 | 22.07 (4.63) | Ref |
| Gargle 1 | 40 | 29.89 (4.34) | < 0.001<sup>a</sup> |
| Gargle 2 | 40 | 29.25 (3.99) | < 0.001<sup>a</sup> |
| Sputum | 16 | 25.82 (9.21) | 0.28<sup>a</sup> |
| Spit | 22 | 23.51 (4.57) | 0.11<sup>a</sup> |
| <sup>a</sup> Paired t-test |  |  |  |
| <sup>b</sup> Wilcoxon signed-rank test |  |  |  |

All patients’ different specimens’ results are presented in supplementary figures 1-4.

## Discussion

We evaluated self-collection of alternative specimens: gargle, spit and sputum and compared their sensitivity and Ct values to NPS specimens as alternative methods for SARS-CoV-2 infection diagnosis. All specimen collection methods were as sensitive as NPS, with sensitivities exceeding 90 %. In comparison with NPS, the gargle specimens had higher Ct values, likely due to dilution by gargling water. Therefore in milder cases with low viral load, it might not be as sensitive as NPS. Spit and sputum specimens collected on the following day had a higher though not significantly different Ct value compared to NPS, however, sputum appeared more challenging to collect in patients with milder symptoms. Hence, spit would appear as the most suitable alternative specimen to NPS for SARS-CoV-2 infection diagnosis.

Since the beginning of the COVID-19 pandemic, several studies or meta-analysis have investigated the potential use of saliva specimen for SARS-CoV-2 infection diagnosis among both symptomatic (8,9,11,16–21) and asymptomatic patients (8,9,11,16–18,21,22). ECDC and CDC have approved the use of saliva or sputum as a diagnostic specimen for COVID-19 for patients with a productive cough (23,24).

There has been mixed results considering difference in Ct levels between NPS and saliva specimens (11,19,20,22). These inconclusive results can be caused by various factors: morning saliva might have higher viral load compared to the rest of the day (18), levels of SARS-CoV-2 virus in saliva correlate to COVID-19 symptoms severity (25). Additionally, early-morning posterior oropharyngeal spitting technique has been considered to have higher sensitivity for SARS-CoV-2 infection diagnosis compared to NPS (8).

Previously, gargle specimens have also been estimated sensitive for SARS-CoV-2 infection diagnosis compared to NPS including 50 inpatients (26) and outpatients (10,27) with confirmed COVID-19. Ct values of gargle specimens were higher than in NPS in these three studies including one with 19 620 outpatients which is in line with findings in our study (10,26,27). Interestingly Goldfard et al, when analyzing 40 COVID-19 outpatients concluded that gargle specimens had higher sensitivity compared to saliva specimens (97.5 % CI 86.8-99.9 % compared to 78.8 CI 95 % 61.0-91.0 %) which is inconsistent with our results, although saliva collection methods were similar (10).

Levican et al and Malcynski et al showed that diagnostic results and Ct values were comparable between sputum and nasopharyngeal tests during the first ten days after COVID-19 diagnosis in study population of 82 and 50 hospitalized patients, respectively (28,29). The result was also consistent with this study.

Alternative specimens’ collection main benefit compared to NPS would be to avoid close contact with a HCW, to allow participants to collect the specimen themselves and to avoid an unpleasant procedure. Overall, it would also increase the willingness to apply for SARS-CoV-2 testing and allocation of current resources in HCWs.

The main limitations of our work are that we did not analyse whether delayed transport or extended storage before analysis could hamper sensitivity as all samples were transported and processed on the very same day and our low sample size (40 participants in total), however we conducted this study as an exploratory assessment of alternative specimen collection. We focused on patients with the most common clinical picture of COVID-19: mild symptoms, as they are the ones that would most benefit from non-invasive alternative specimen collection. An additional strength was that all samples were collected within 1-2 days after diagnosis, while patients were still at the acute phase of the disease.

## Conclusion

Among gargle, spit and sputum specimens, morning collection of approximately 30 mL of spit before any food and water intake or teeth brushing appeared to be the most suitable alternative specimen collection method with a Ct-value as low as obtained with NPS and ease of collection for patients with mild symptoms. Our findings were promising but warrant further investigations with larger study population. We will consider offering the possibility to patients with mild symptoms seeking diagnosis in a pilot testing centre, the possibility to enroll in a study assessing whether in general population, spit collected on the next morning has the same performance as in our exploratory sample. If so, in the long run, we could offer patients with mild symptoms to choose between at home self-collection of spit versus nasopharyngeal specimen collection at a testing centre. Not only would it decrease discomfort, but also decrease HCWs’ exposure and burden.

## Data Availability

Data is available anonymously

## Acknowledgements

The authors thank Helsinki city and Helsinki city’s Epidemiological Unit for sharing data of the COVID-19 patients. We are also grateful to THL Virology laboratory staff who analyzed the specimens among other works.

## Author’s contributions

Study design: Henna Mäkelä, Lotta Hagberg, Thuan Vo, Hanna Nohynek, Niina Ikonen, Otto Helve, Carita Savolainen-Kopra and Timothée Dub.

Statistical analysis: Thuan Vo

Specimen collection and logistics: Eero Poukka, Henna Mäkelä, Lotta Hagberg and Timothée Dub.

Laboratory analysis: Niina Ikonen, Kirsi Liitsola and THL Virology laboratory.

Writing and editing: All authors.

## Tables and figures

**Attachment 1.**
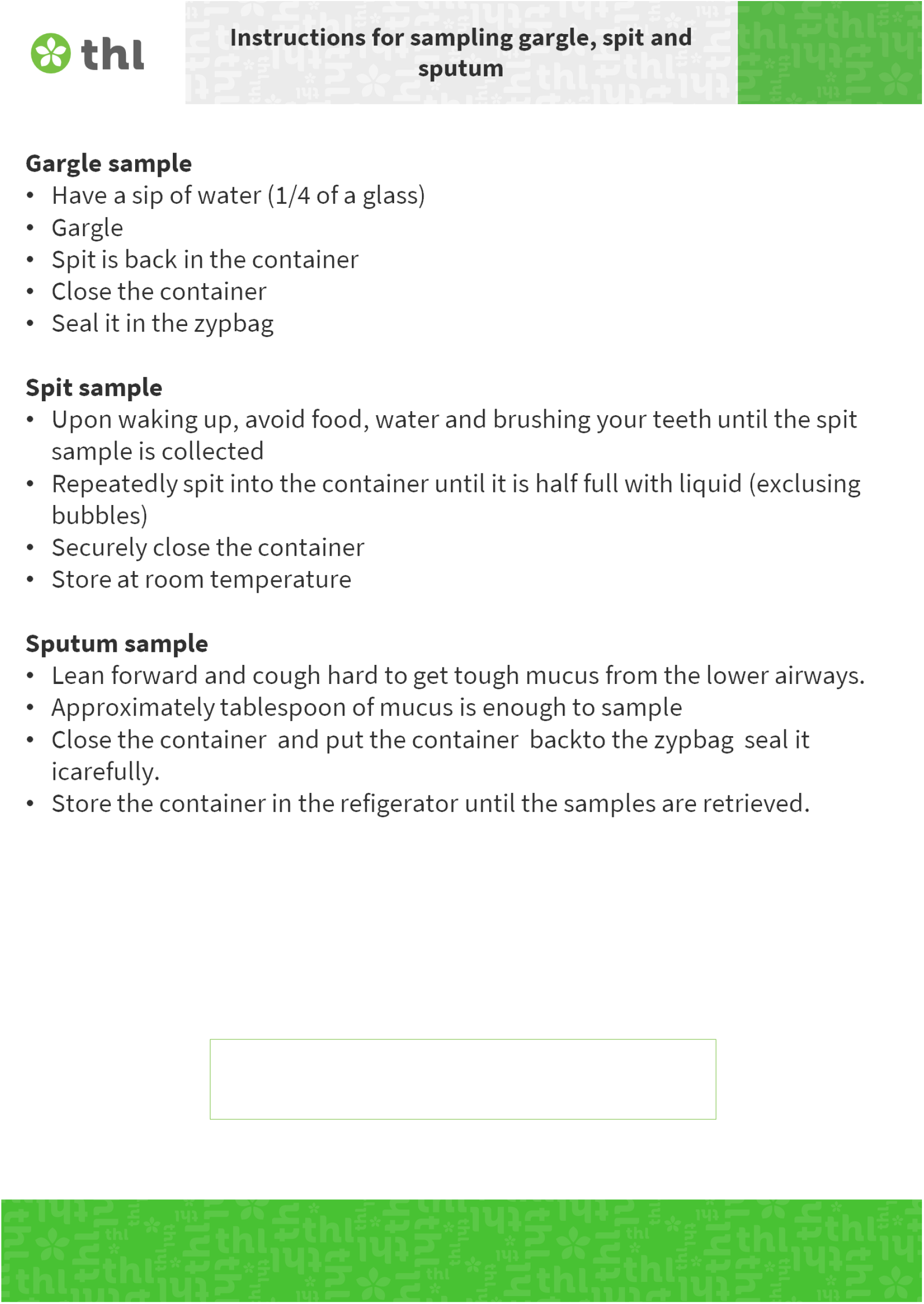
Instructions for sampling gargle, spit and sputum.

**Suplementary table 1.**
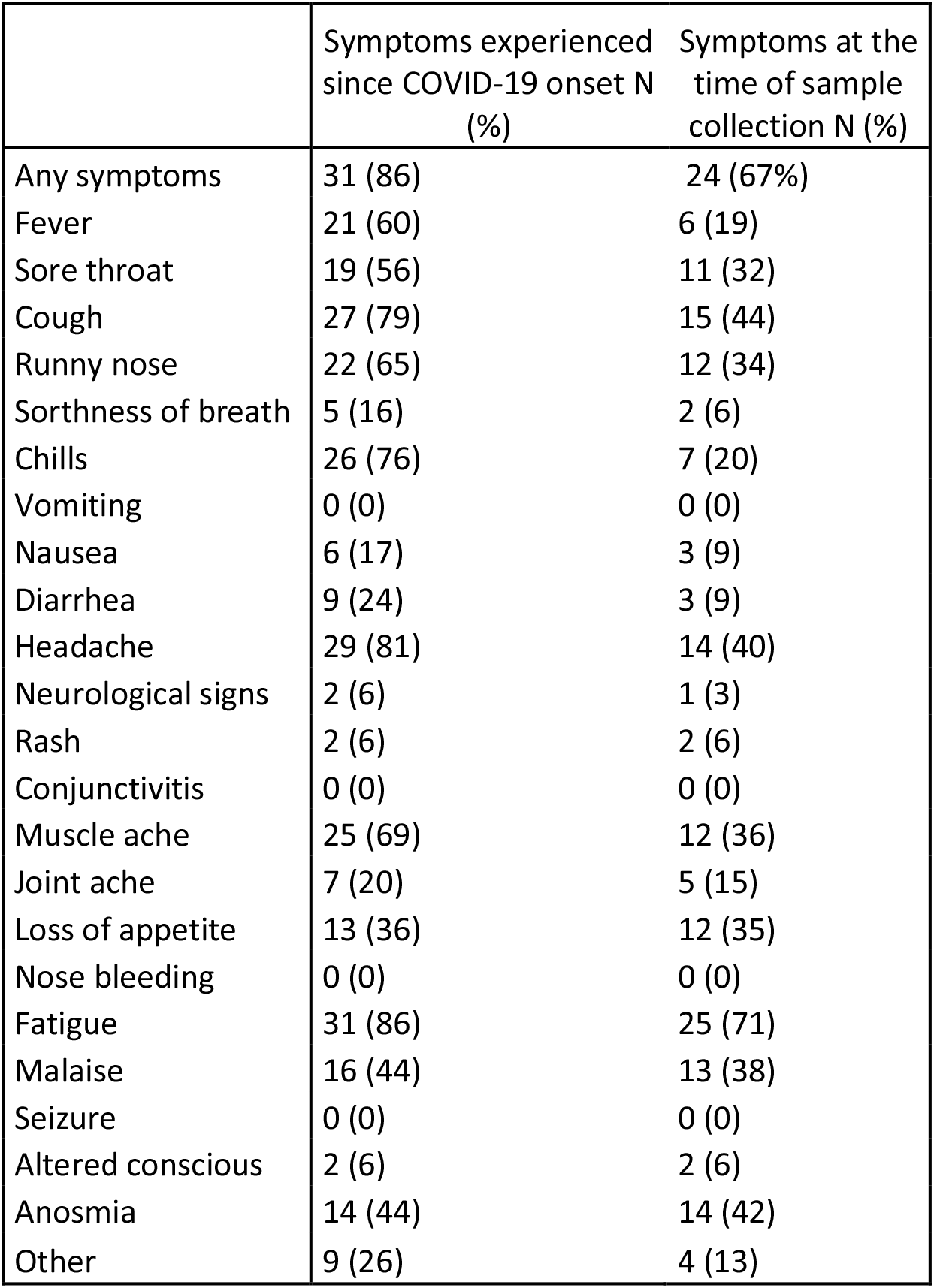
Symptoms reported by patients.

